# Older biological age is associated with adverse COVID-19 outcomes: A cohort study in UK Biobank

**DOI:** 10.1101/2021.03.20.21254010

**Authors:** Qingning Wang, Veryan Codd, Zahra Raisi-Estabragh, Crispin Musicha, Vasiliki Bountziouka, Stephen Kaptoge, Elias Allara, Emanuele Di Angelantonio, Adam S. Butterworth, Angela M. Wood, John R. Thompson, Steffen E Petersen, Nicholas C. Harvey, John N. Danesh, Nilesh J. Samani, Christopher P. Nelson

## Abstract

**Background:** Older chronological age is the most powerful risk factor for adverse coronavirus disease-19 (COVID-19) outcomes. It is uncertain, however, whether older biological age, as assessed by leucocyte telomere length (LTL), is also associated with COVID-19 outcomes.

**Methods:** We associated LTL values obtained from participants recruited into UK Biobank (UKB) during 2006-2010 with adverse COVID-19 outcomes recorded by 30 November 2020, defined as a composite of any of the following: hospital admission, need for critical care, respiratory support, or mortality. Using information on 131 LTL-associated genetic variants, we conducted exploratory Mendelian randomisation (MR) analyses in UKB to evaluate whether observational associations might reflect cause-and-effect relationships.

**Findings:** Of 6,775 participants in UKB who had tested positive for infection with SARS-CoV-2 in the community, there were 914 (13.5%) with adverse COVID-19 outcomes. The odds ratio (OR) for adverse COVID-19 outcomes was 1·17 (95% CI 1·05-1·31; P=0·004) per 1-SD shorter usual LTL, after adjustment for chronological age, sex and ethnicity. Similar ORs were observed in analyses that: adjusted for additional risk factors; disaggregated the composite outcome and reduced the scope for selection or collider bias. In MR analyses, the OR for adverse COVID-19 outcomes was directionally concordant but non-significant.

**Interpretation:** Shorter LTL, indicative of older biological age, is associated with higher risk of adverse COVID-19 outcomes, independent of several major risk factors for COVID-19 including chronological age. Further data are needed to determine whether this association reflects causality.

**Funding:** UK Medical Research Council, Biotechnology and Biological Sciences Research Council and British Heart Foundation.

## Introduction

Older chronological age has emerged as the most powerful risk factor for severe infection, requiring hospitalisation or critical care, and mortality from coronavirus disease 19 (COVID-19) caused by severe acute respiratory syndrome coronavirus 2 (SARS-CoV-2).^1,2^ One potential mediator of this effect is ageing of the immune system, leading to increased levels of pro-inflammatory senescent cells and reduced proliferative capacity of immune precursor cells.^3,4^ Whether an individual’s biological age, which may be older or younger than their chronological age, plays an additional role in COVID-19 outcome is unclear. Telomere length (TL), a proposed marker of biological age, is a key determinant of proliferative capacity and cellular lifespan, triggering senescence once a critically short TL is reached.^5^ Although TL − commonly measured in leucocytes (LTL) − declines with chronological age, age accounts for only ∼3.5% of the inter-individual variation in LTL.^6^

A few small case-control studies, in which LTL was measured after SARS-CoV-2 infection at the time of hospital admission, have reported associations of shorter LTL with hospitalisation and severe outcomes.^7−9^ However, their interpretation is complicated by the possibility that LTL measurements could have been influenced by white cell turnover in response to infection. To our knowledge, no study to date has reported on associations of prior (pre-infection) LTL values and adverse COVID-19 outcomes.

Here, we examine whether LTL measured several years prior to SARS-CoV-2 infection is associated with adverse COVID-19 outcomes, leveraging our recent completion of LTL measurements in 474,074 participants aged 40-69 at time of recruitment into UK Biobank (UKB)^6^ between 2006 and 2010.^10, 11^

## Methods

### Participants

Participants in UKB have been characterised in detail using questionnaires, physical measurements, urinary and plasma biomarker measurements, genomic assays and longitudinal linkage with multiple health record systems, including Hospital Episode Statistics (HES) and Office for National Statistics (ONS) mortality data.^12^ We have described the associations of inter-individual variation in LTL with multiple biomedical traits and risk of several diseases in UKB.^11^ Since the onset of the COVID-19 pandemic, UKB has also linked participants with results from clinically indicated SARS-CoV-2 testing and COVID-19 outcomes. By linking participants in UKB to SARS-CoV-2 testing datasets of Public Health England (PHE),^13^ we identified participants who tested positive between 16 March 2020 and 30 November 2020; the latter date corresponds to the latest release of HES data to UKB. We used HES records to identify SARS-CoV-2 positive participants who were admitted to hospital due to COVID-19 (ICD-10 code ‘U07.1’) within 28 days after a positive SARS-CoV-2 test. We further extracted information on need for critical care admission and respiratory support, due to COVID-19 (ICD-10 code ‘U07.1’), via linkage to the ICNARC (Intensive Care National Audit and Research Centre) database, and deaths due to COVID-19 (ICD-10 code ‘U07.1’), from the Office for National Statistics (ONS) death registry data.

The UK Biobank has ethical approval from the North West Centre for Research Ethics Committee (Application 11/NW/0382), which covers the UK. UK Biobank obtained informed consent from all participants. Full details can be found at https://www.ukbiobank.ac.uk/learn-more-about-uk-biobank/about-us/ethics. The work in this manuscript was approved by UK Biobank (Application 6077). The generation and use of the data presented in this paper was approved by the UK Biobank access committee under UK Biobank application number 6077.

### LTL measurements

Full details of the LTL measurements in UKB are provided elsewhere.^6^ Briefly, LTL was measured using a validated PCR method that expresses LTL as a ratio (T/S ratio).^6^ LTL measurements were adjusted for technical variation, log_e_ transformed and Z-standardised.^6^ We made paired LTL measurements at two time-points (mean interval: 5·5 years) in 1,351 participants, yielding a regression-dilution ratio of ∼0·68. Results in this study have been corrected for within-person variability of LTL values over time (abbreviated “usual LTL”), as described previously.^6,11^

### Primary outcome

Our study’s primary outcome was a composite of COVID-19-related outcomes (ICD-10 code ‘U07.1’): hospital admission, requirement for critical care, respiratory support, or mortality. We defined cases as those participants in UKB who tested positive for SARS-CoV-2 and had the primary outcome. Controls were those who tested positive for SARS-CoV-2 but did not have the primary outcome. To reduce the scope for collider bias^14^ we included only participants with positive SARS-CoV-2 tests done outside of hospital settings, since hospital admission itself may increase the likelihood of SARS-CoV-2 testing. The chronological age, sex and ethnicity adjusted odds ratio (OR) for having a SARS-CoV-2 test (n=43,574) at any location, was 1·03, (95% CI 1·01-1·05; P=1·0×10^−4^) per 1-SD shorter usual LTL.

### Statistical analysis

Analyses involved multivariable logistic regression, adjusting for chronological age (at SARS-CoV-2 positive test), sex and ethnicity. Due to small numbers, ethnic groups other than White were combined and participants with missing ethnicity (n=14 cases and 46 controls) were excluded. To remove the correlation between LTL and chronological age, we used the residuals of LTL adjusted for chronological age at baseline within the statistical models. ORs were further adjusted for baseline smoking status and body-mass index (BMI) recorded at entry into UKB. Results are described as ORs associated with the outcome per one standard deviation (SD) shorter LTL residual, with associated 95% confidence intervals (CI) and p-values.

We conducted several secondary analyses. First, we examined associations with each component of the primary composite endpoint. Second, we analysed the primary outcome using the rest of UKB participants as controls, as testing was unlikely to be random and the restriction to SARS-CoV-2 positive controls only is potentially subject to selection bias related to factors associated with infection.^15^ Third, to ensure that apparently post-COVID-19 outcomes were not re-admissions or influenced by proximate medical events prior to infection, we excluded participants with any hospital admission in the previous 6 months. Fourth, to consider the impact of baseline disease prevalence on LTL, and to minimise the potential confounding by these diseases on any LTL-COVID-19 outcomes relationship, we used age- and disease-adjusted LTL residuals, based on 123 previously defined diseases.^11^ Finally, in an exploratory analysis, we conducted one-sample Mendelian randomisation (MR) analyses in UKB to evaluate a causal relationship between shorter LTL and adverse COVID-19 outcomes, using the inverse-variance weighted (IVW)^16^ and weighted median^17^ methods with a set of 131 independent and uncorrelated genetic variants we recently identified for LTL.^11^ We used MR-Egger regression to assess robustness to horizontal pleiotropy.^18^

## Results

By 30 November 2020, 914 participants were identified with an adverse COVID-19 related outcome and 5861 participants were identified as primary controls (positive community test for COVID-19 but not hospitalised). Their characteristics are summarised in **Table 1**. On average, compared to controls, cases were older and more likely to be male and from a non-White background. At time of their entry into UKB, they also had a higher BMI and more likely to be current smokers. (**Table 1**).

**Table 1.**
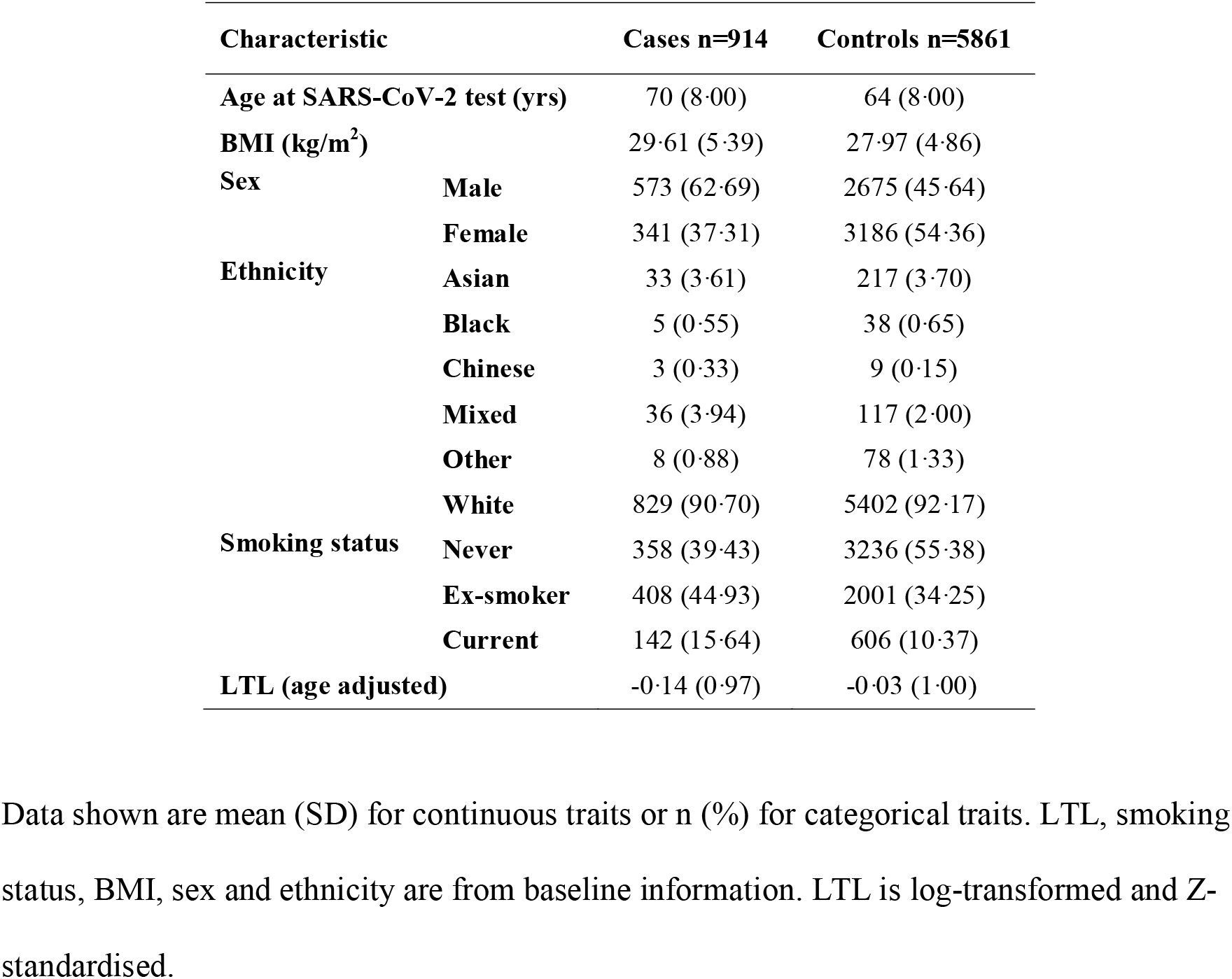
Characteristics of participants by case status.

LTL at entry to UKB was on average shorter in cases compared with controls (**Table 1**). The OR for the primary outcome was 1.17 (95% CI 1.05-1.31; P=0·004) per 1-SD shorter usual LTL, after adjustment for chronological age, sex and ethnicity (**Table 2**). The OR only slightly attenuated after further adjustment for smoking status and BMI (OR=1·15, 1·03-1·28), and after adjustment for the presence of any of 123 diseases recorded at baseline (OR=1·14, 1·02-1·27). As expected, older chronological age, male sex and non-White ethnicity were each associated with higher risk of adverse COVID-19 outcomes independently of usual LTL (**Table 2**).

**Table 2.** Results of the main and secondary/sensitivity analyses.

|  | N cases | N controls | Odds Ratio (95%CI) | P-value |
| --- | --- | --- | --- | --- |
| **Composite outcome** |  |  |  |  |
| LTL (age-adjusted) (per 1 SD shorter) |  |  | 1.17 (1.05 , 1.31) | 0.004 |
| Age at COVID-19 test (per 5 yrs older) | 914 | 5861 | 1.58 (1.51 , 1.65) | <0.001 |
| Sex (male vs female) |  |  | 1.88 (1.62 , 2.19) | <0.001 |
| Ethnicity (non-White vs White) |  |  | 1.80 (1.39 , 2.34) | <0.001 |
| **Separate components as outcome\*** |  |  |  |  |
| Hospitalisation | 672 |  | 1.17 (1.03 , 1.32) | 0.013 |
| Critical care support | 383 | 5861 | 1.31 (1.12 , 1.54) | <0.001 |
| Respiratory support | 279 |  | 1.36 (1.13 , 1.64) | <0.001 |
| Death | 157 |  | 1.36 (1.07 , 1.73) | 0.013 |
| **Population controls** |  |  |  |  |
| LTL (age-adjusted) (per 1 SD shorter) | 914 | 465,946 | 1.19 (1.08 , 1.31) | <0.001 |
| **Excluding participants with recent hospitalisation** |  |  |  |  |
| LTL (age-adjusted) (per 1 SD shorter) | 732 | 5861 | 1.15 (1.02 , 1.30) | 0.019 |
| **Mendelian Randomisation** |  |  |  |  |
| MR IVW | 914 | 5861 | 1.48 (0.79 , 2.77) | 0.224 |
| MR-median |  |  | 1.38 (0.50 , 3.86) | 0.537 |

The main analysis is based on our composite outcome and the full multivariable model estimates are shown for each risk factor. \*For each component of the composite outcome analysed separately, the results shown for these are labelled by outcome component but represent the LTL (age-adjusted) estimate (per 1 SD shorter). For each analysis, the numbers of cases and controls are given alongside the odds ratio, 95% confidence interval and P-value. MR IVW: Mendelian randomisation inverse-variance weighted method. MR-median: Mendelian randomisation weighted median method.

Sub-components of our study’s primary composite outcome were not mutually exclusive, as 46 cases contributed to all four sub-components (**Figure 1**). Shorter usual LTL was significantly associated with higher risk of each sub-component (**Table 2**). ORs were broadly similar to the main findings in analyses that replaced the SARS-CoV-2-positive control group with all UKB participants as controls or that excluded any participant with a hospital admission in the six months prior to testing positive for SARS-CoV-2 (**Table 2**).

**Figure 1.**
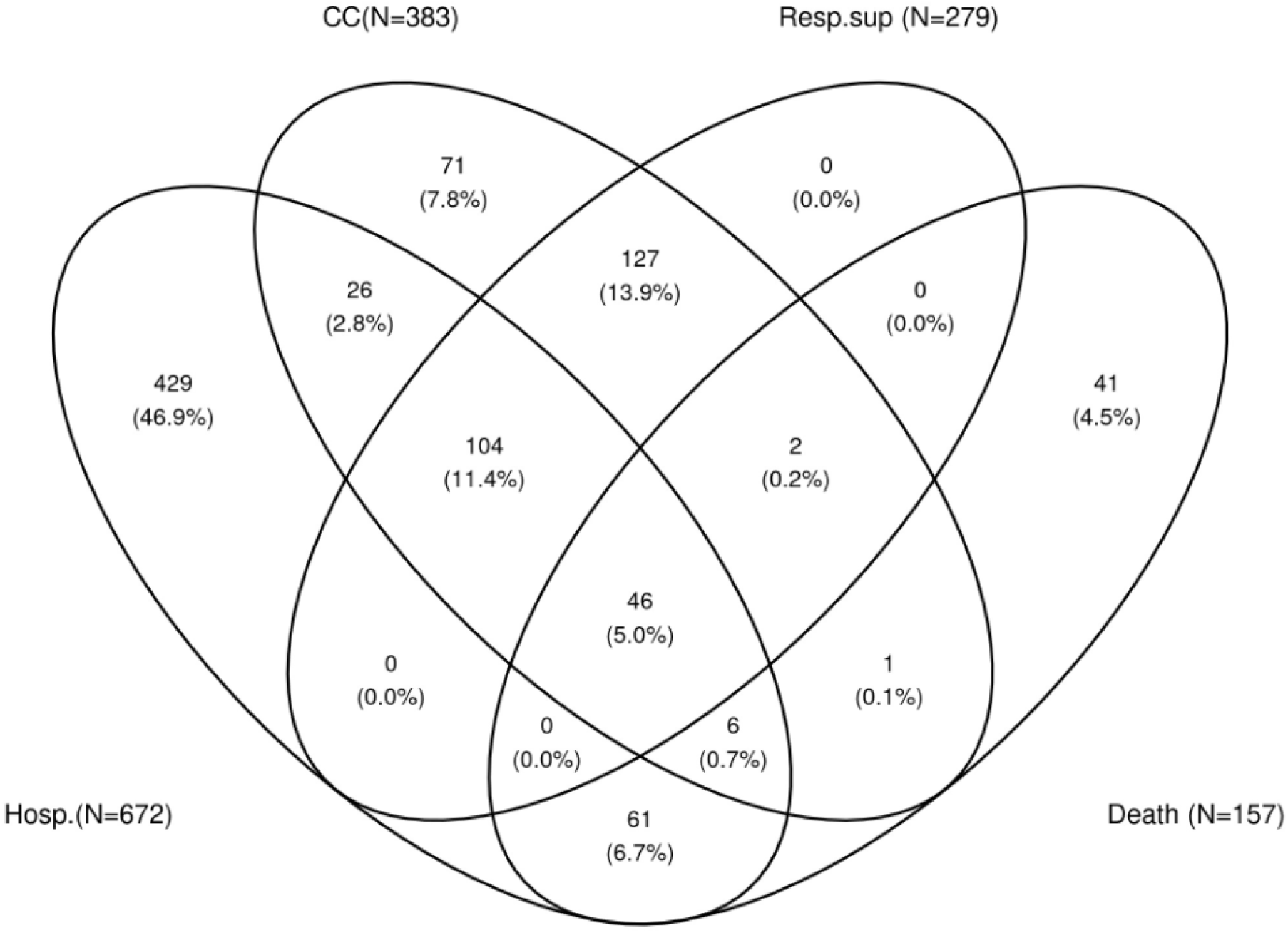
Venn diagram showing the distribution of the individual components of the primary outcome. Hosp., hospitalised due to COVID-19; CC, critical care admission due to COVID-19; Resp. sup, need for respiratory support; Death, death due to COVID-19.

In MR analyses, the IVW odds ratio was 1·48 (0·79-2·77; P=0·224) per 1-SD shorter genetically-determined LTL, a non-significant result directionally concordant with the observational finding (**Table 2**). Results were similar using the weighted median method (**Table 2**) and there was no evidence of horizontal pleiotropy (MR-Egger intercept P=0·591).

## Discussion

In a study of 6,775 participants with a positive test for SARS-CoV-2 (nested within the 500,000-participant UKB), we have shown that individuals with shorter LTL assessed several years *prior* to SARS-CoV-2 infection had higher risk of adverse COVID-19 outcomes, even after adjustment for several established risk factors for COVID-19 including chronological age. This finding suggests that being “biologically” older is likely to be independently associated with COVID-19 hospitalisation and severity. The results of analysis of LTL-associated genetic variants and COVID-19 were directionally concordant with our observational findings but non-significant. Our results, therefore, encourage further investigation of the potential causal relevance of biological ageing to adverse COVID-19 outcomes.

The validity of our results is supported by several observations. First, our study confirmed well-known associations of older chronological age, male sex, and non-White ethnicity with adverse COVID-19 outcomes.^2^ Each of these factors was associated much more strongly with COVID-19 outcomes than was shorter LTL. Second, we found significant associations of shorter LTL with each sub-component of our study’s primary composite outcome. Third, our main findings persisted after adjustment for multiple risk factors. Fourth, our overall result was robust to sensitivity analyses designed to minimise the scope for potential biases. For example, collider bias can lead to false associations between a risk factor and an outcome,^14^ as highlighted by studies related to understanding of COVID-19 disease risk and severity.^15^ Indeed, we found evidence for potential colliders in our own analysis, observing a small but significant association between shorter LTL and higher likelihood of SARS-CoV-2 testing. Hence, we only included participants with a positive SARS-CoV-2 test outside the hospital setting, as hospitalisation itself may increase the likelihood of testing.

The biological mechanisms through which shorter LTL might increase risk of adverse outcomes from SARS-CoV-2 infection remain to be clarified. Our finding that the association was not substantially attenuated when we adjusted for the association of LTL with multiple diseases at baseline, suggest that, if this association is causal,, it is probably not simply a reflection of co-morbidity due to the impact of shorter LTL on risk of these diseases. A potential mechanism relates to the impact of telomere length dynamics on aging of the immune system^19^ and the potential role of senescence in severe SARS-CoV-2 infection.^3,4,20^ When challenged with infection, individuals with shorter LTL prior to infection would have less proliferative capacity and be more likely to accrue high levels of senescent cells more quickly than individuals with longer LTL.^21^ Individuals with shorter LTL may therefore potentially already harbour a higher proportion of senescent T-cells, reducing the number of functional cells that are able to respond to infection.^20^ Additionally, senescent cells are known to adopt a pro-inflammatory phenotype, secreting high levels of cytokines, which can further drive inflammation in COVID-19 patients.^20^

Our study has several limitations. UKB is not representative of the general UK population; only 6% of those invited to participate did so.^22^ Risk factor levels and mortality rates are lower than in the general population, although risk factor associations with mortality for a range of diseases are similar.^23^ Hence, further studies are warranted in other populations. Our one-sample Mendelian randomisation analysis in UKB had limited power to reliably estimate causal effects as fewer than one thousand participants had been hospitalised after a positive SARS-CoV-2 test and our genetic instrument of 131 variants, while using the most up to date information on LTL-associated variants, accounts for only ∼4% of inter-individual variation in LTL.^11^ While there are data from large genetic studies of COVID-19^24^, they could not be used in our analysis because the outcome definitions differed substantially from those we used, and because of their inclusion of within hospital testing that is potentially a collider with LTL and COVID-19 outcomes. Larger sample sizes with comparable disease phenotypes should, therefore, enable more precise evaluation of a potential causal association between shorter LTL and adverse COVID-19 outcomes.

In conclusion, in the largest study to date, we provide evidence that shorter LTL, reflecting older biological age, is associated with higher risk of adverse COVID-19 outcomes, independent of several major risk factors for COVID-19.

## Data Availability

Source data is accessible via application to the UK Biobank.

## Data sharing

Source data is accessible via application to the UK Biobank.

## Acknowledgements

This research has been conducted using the UK Biobank Resource under Application Number 6077 and was funded by the UK Medical Research Council (MRC), Biotechnology and Biological Sciences Research Council and British Heart Foundation (BHF) through MRC grant MR/M012816/1. C.P.N is funded by the BHF (SP/16/4/32697). V.C., C.M., V.B., Q.W., C.P.N. and N.J.S. are supported by the National Institute for Health Research (NIHR) Leicester Cardiovascular Biomedical Research Centre (BRC-1215-20010). Cambridge University investigators are supported by the B.H.F (RG/13/13/30194; RG/18/13/33946), Health Data Research UK, NIHR Cambridge Biomedical Research Centre (BRC-1215-20014), NIHR Blood and Transplant Research Unit in Donor Health and Genomics (NIHR BTRU-2014-10024) and MRC (MR/L003120/1). J.D. holds a BHF Personal Professorship and NIHR Senior Investigator Award. A.M.W. and E.A. received support from the EU/EFPIA Innovative Medicines Initiative Joint Undertaking BigData@Heart (11607). Z.R.E. is supported by BHF Clinical Research Training Fellowship No. FS/17/81/33318. S.E.P. acknowledges support from the NIHR Barts Biomedical Research Centre.

## Author Contributions

V.C., C.P.N., J.N.D., S.E.P., N.C.H. and N.J.S. conceived the project. All authors contributed to the sample definition and the analysis plan. Q.W., C.M. and C.P.N. performed the analyses. V.C., C.P.N., Q.W. and N.J.S. prepared the manuscript and all authors revised it. V.C., C.P.N., J.R.T., J.N.D. and N.J.S. (Principal investigator) secured funding and oversaw the project.

## Competing Interests Declaration

The authors declare no competing interests.

Data shown are mean (SD) for continuous traits or n (%) for categorical traits. LTL, smoking status, BMI, sex and ethnicity are from baseline information. LTL is log-transformed and Z-standardised.

